# The effect of herpes zoster vaccines on incident dementia in England: Protocol for an OpenSAFELY regression discontinuity study

**DOI:** 10.64898/2026.09.03.26362052

**Authors:** Venexia Walker, Andrea Schaffer, Will Hulme, Christopher Wood, Arina Tamborska, Alex Walker, Brian MacKenna, Ben Goldacre, Jonathan Sterne, Pascal Geldsetzer

**Author notes:** Contributed equally (Ben Goldacre, Jonathan Sterne, Pascal Geldsetzer).

## Abstract

The English national shingles vaccination programme was officially launched on 1 September 2013 using the live attenuated herpes zoster vaccine Zostavax and updated on 1 September 2023 to replace Zostavax with the two-dose, non-live, recombinant subunit herpes zoster vaccine Shingrix. A study using a regression discontinuity design, conducted following the introduction of the herpes zoster vaccination programme on 1 September 2013 in Wales and using the SAIL databank to analyse Welsh routinely collected data, suggested that Zostavax vaccination reduced the probability of a new dementia diagnosis by approximately 20% over a seven-year follow-up period. The effects of Zostavax and Shingrix on incident dementia in England have not been published to date. This protocol describes a regression discontinuity study to study the effect of herpes zoster vaccines on incident dementia in England using linked electronic health records from OpenSAFELY.

## Background

The English national shingles vaccination programme was officially launched on 1 September 2013 using the live attenuated herpes zoster (HZ) vaccine Zostavax. In the first year of the programme, the vaccine was routinely offered to adults aged 70 years on 1 September 2013 (i.e., born between 2 September 1942 and 1 September 1943) and to adults aged 79 on 1 September 2013 (i.e., born between 2 September 1933 and 1 September 1934) as part of the catch-up campaign. (1) Over time, the programme was updated (**Table 1**) and on 1 April 2017 it was simplified so that adults became eligible on their 70th birthday (routine cohort) or their 78th birthday (catch-up cohort). In September 2020, the catch-up programme was completed.

**Table 1:**
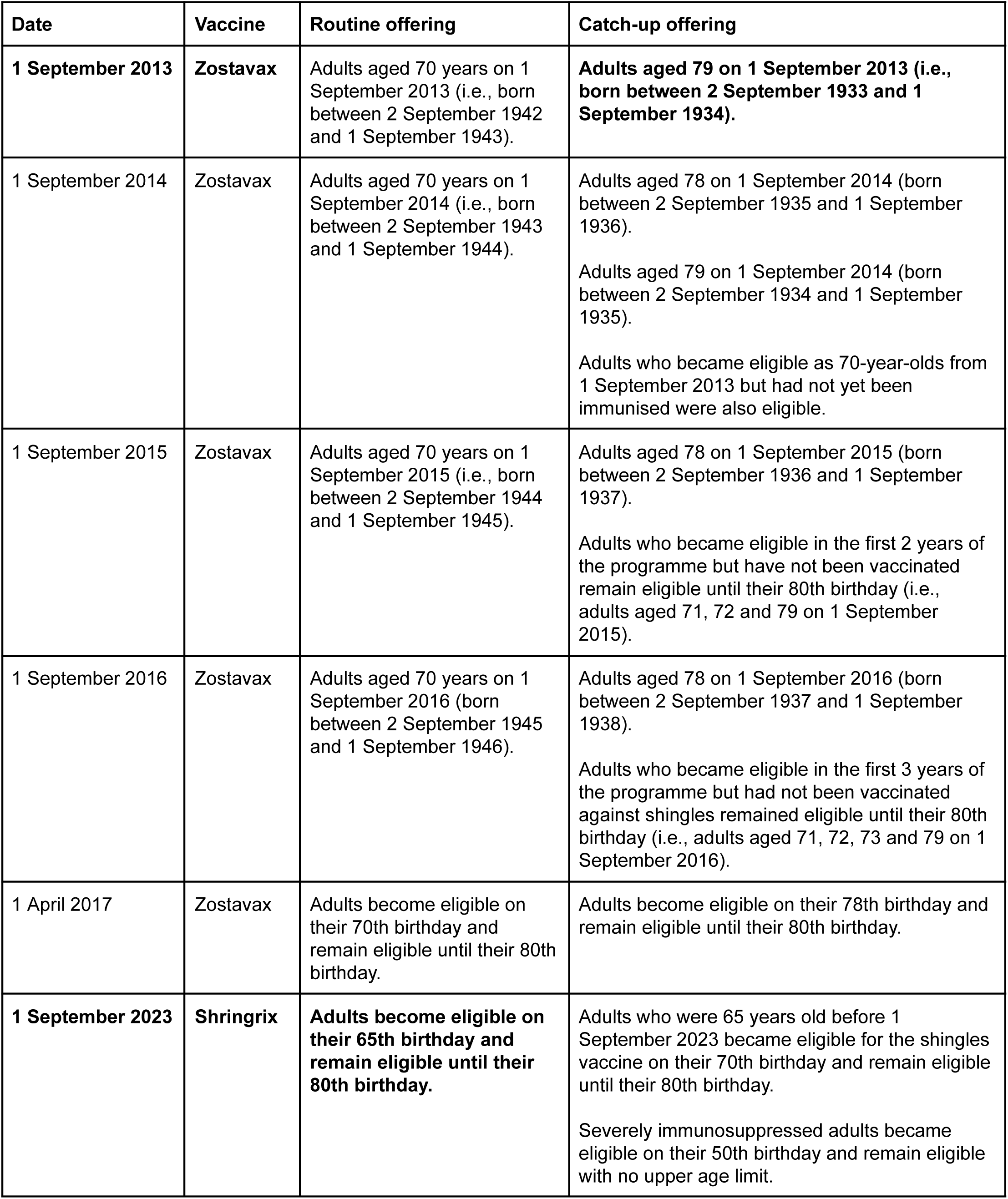
Routine and catch-up offerings for the Shingles vaccine in England from 2013 to 2023. Derived from: <u>Shingrix vaccine update report</u>. Bold text represents the programmes evaluated for aims 1 (Zostavax) and 2 (Shingrix).

| Date | Vaccine | Routine offering | Catch-up offering |
| --- | --- | --- | --- |
| **1 September 2013** | **Zostavax** | Adults aged 70 years on 1 September 2013 (i.e., born between 2 September 1942 and 1 September 1943). | **Adults aged 79 on 1 September 2013 (i.e., born between 2 September 1933 and 1 September 1934).** |
| 1 September 2014 | Zostavax | Adults aged 70 years on 1 September 2014 (i.e., born between 2 September 1943 and 1 September 1944). | Adults aged 78 on 1 September 2014 (born between 2 September 1935 and 1 September 1936).<br>Adults aged 79 on 1 September 2014 (born between 2 September 1934 and 1 September 1935).<br>Adults who became eligible as 70-year-olds from 1 September 2013 but had not yet been immunised were also eligible. |
| 1 September 2015 | Zostavax | Adults aged 70 years on 1 September 2015 (i.e., born between 2 September 1944 and 1 September 1945). | Adults aged 78 on 1 September 2015 (born between 2 September 1936 and 1 September 1937).<br>Adults who became eligible in the first 2 years of the programme but have not been vaccinated remain eligible until their 80th birthday (i.e., adults aged 71, 72 and 79 on 1 September 2015). |
| 1 September 2016 | Zostavax | Adults aged 70 years on 1 September 2016 (born between 2 September 1945 and 1 September 1946). | Adults aged 78 on 1 September 2016 (born between 2 September 1937 and 1 September 1938).<br>Adults who became eligible in the first 3 years of the programme but had not been vaccinated against shingles remained eligible until their 80th birthday (i.e., adults aged 71, 72, 73 and 79 on 1 September 2016). |
| 1 April 2017 | Zostavax | Adults become eligible on their 70th birthday and remain eligible until their 80th birthday. | Adults become eligible on their 78th birthday and remain eligible until their 80th birthday. |
| **1 September 2023** | **Shingrix** | **Adults become eligible on their 65th birthday and remain eligible until their 80th birthday.** | Adults who were 65 years old before 1 September 2023 became eligible for the shingles vaccine on their 70th birthday and remain eligible until their 80th birthday.<br>Severely immunosuppressed adults became eligible on their 50th birthday and remain eligible with no upper age limit. |

The English national shingles vaccination programme was updated on 1 September 2023 to replace Zostavax with the two-dose, non-live, recombinant subunit HZ vaccine Shingrix (**Table 1**). From 1 September 2023, the Shingrix vaccine was routinely offered to adults on their 65th birthday; while adults who were 65 years old before 1 September 2023 became eligible for the shingles vaccine on their 70th birthday. People remained eligible to receive their first dose until their 80th birthday and their second dose until their 81st birthday. In addition, severely immunosuppressed adults became eligible for the Shingrix vaccine on their 50th birthday with no upper age limit, due to their increased risk of HZ.

A study using a regression discontinuity (RD) design, conducted following the introduction of the HZ vaccination programme on 1 September 2013 in Wales and using the SAIL databank to analyse Welsh routinely collected data, suggested that Zostavax vaccination reduced the probability of a new dementia diagnosis by approximately 20% over a seven-year follow-up period. (2) The effects of Zostavax and Shingrix on incident dementia in England have not been published to date.

### Objectives

The objective of the study is to estimate the effects of Zostavax and Shingrix vaccination on incident dementia in England. Specifically, we aim to:

1. Estimate the effect of eligibility versus non-eligibility for Zostavax vaccination on 1 September 2013 on incident dementia
2. Estimate the effect of eligibility versus non-eligibility for Shingrix vaccination at age 65 on incident dementia

We will explore the feasibility of estimating the effect of eligibility versus non-eligibility for Shingrix vaccination on your 70th birthday on incident dementia. This analysis requires further consideration due to the phased transition between the Zostavax programme and the Shingrix programme, which meant some adults aged 70 to 80 years could receive either vaccine depending on timing, stock availability, and clinical status. We will publish a protocol amendment to pursue this aim if it proves feasible.

For aim 1, we aim to replicate as closely as possible the regression discontinuity analysis conducted following the introduction of the HZ vaccination programme for Zostavax on 1 September 2013 in Wales, with analytic choices prespecified as much as possible. (2) Deviations from the Welsh analysis are summarised in **Supplementary Table 1**.

## Methods

### OpenSAFELY platform

All data will be linked, stored and analysed securely using the OpenSAFELY platform, https://www.opensafely.org/, as part of the NHS England OpenSAFELY Analytics Service. Primary care records managed by the GP software provider, TPP, have been linked to hospital admissions and ONS registered deaths through OpenSAFELY. No data from patients who registered a Type-1 Opt Out with their GP surgery are included in this study.

The analysis datasets will be defined and created using ehrQL and Python 3.10.12. Analysis will be executed using R 4.4.3. All analysis code will be shared openly for review and re-use under MIT open license on GitHub. The public activity log for this project and a link to the GitHub can be found at https://jobs.opensafely.org/the-effect-of-herpes-zoster-vaccines-on-incident-dementia-in-england/. All future iterations of the pre-specified study protocol will be archived with version control on GitHub: https://github.com/opensafely/zostavax-dementia.

### Study design

Aim 1 considers the Zostavax catch-up offering, under which adults aged 79 on 1 September 2013 (i.e., born between 2 September 1933 and 1 September 1934) were offered Zostavax, but those born on or before 31 August 1933 were never offered Zostavax. We will use an RD study design with calendar time as the ‘running variable’ (the variable that determines whether someone receives treatment, based on whether an individual falls above or below a threshold). This exploits the discontinuity in eligibility on 1 September 2013. The running variable will be:

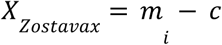

where, for person *i*, *m* is their month of birth (continuous), and *c* is the month of the threshold birth date

Aim 2 considers the Shingrix routine offering, under which adults were offered Shingrix on their 65th birthday and remained eligible until their 80th birthday, but those not yet 65 years of age were not offered Shingrix. We will use an RD study design with age as the running variable. This exploits the discontinuity where adults who turned 65 years old on or after 1 September 2023 (i.e., born on or after 1 September 1958) were eligible for Shringrix, but adults who were 65 years old before 1 September 2023 were not eligible for Shringrix until they turned 70 years old. The running variable will be:

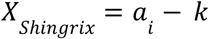

where, for person i, a is their age (continuous; months), and k is the age threshold for eligibility

In both cases, threshold-based eligibility approximates randomisation, as confounding variables are expected to be similar among people just above and below the threshold date.

### Key dates

The threshold date for aim 1 (Zostavax) is 1 September 2013 and for aim 2 (Shingrix) is 1 September 2023. For aim 1, the index date (the date when study follow-up starts) will be 1 February 2014, by which time we expect that approximately 50% of eligible people are vaccinated (**Figure 1**). (3) For aim 2, the index date will be decided following preliminary analyses, described below, that do not include any outcome data. Note that, although age is the running variable in the RD study design for aim 2 (Shingrix), the index date will be the common calendar date corresponding to the plateauing of Shingrix coverage among newly eligible adults (**Figure 2**).

**Figure 1:**
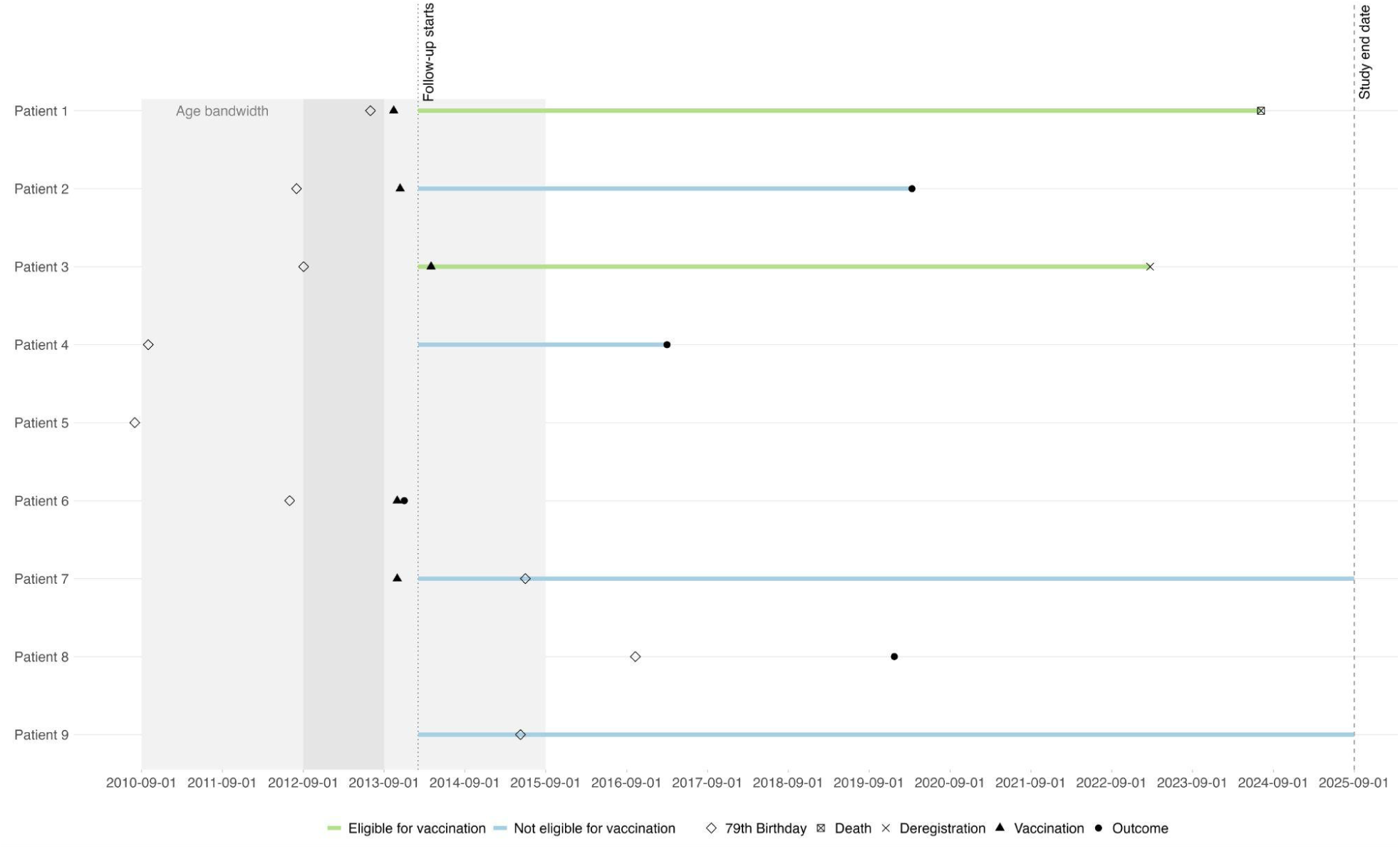
Swimmer plot demonstrating potential patient pathways for aim 1 (Zostavax)

**Figure 2:**
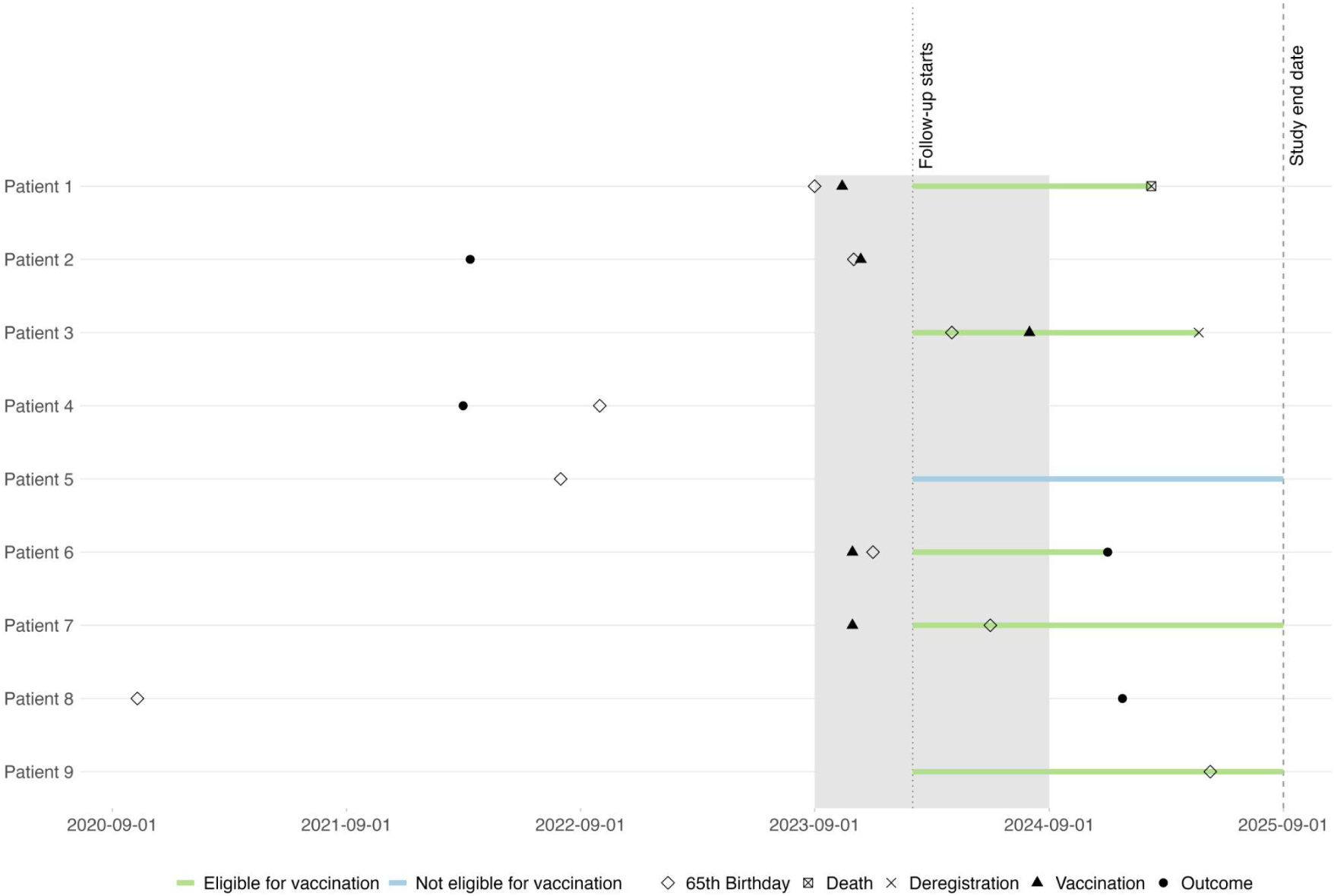
Swimmer plot demonstrating potential patient pathways for aim 2 (Shingrix)

### Data sources

#### Primary care data from TPP

SystmOne is a primary care clinical information system run by TPP, used by roughly one third of GP practices in England, with records for approximately 44% of the English population. (4,5) It captures information about a patient that is electronically recorded or accessed by GPs, including symptoms, investigations, test results, diagnoses, prescriptions, demographic and social characteristics.

#### Hospital admissions

Admitted Patient Care (APC) is the national data set for hospital admissions. Admitted Patient Care Spells (APCS) is part of Hospital Episode Statistics (HES) and is provided to OpenSAFELY via NHS Digital’s Secondary Uses Service (SUS). It includes admission and discharge dates; routes of admission; reason for admission; diagnoses; treatments; and discharge destination. Data is available from early 2016 onwards.

#### ONS registered deaths

Date and cause of death based on information recorded when deaths are certified and registered in England and Wales. The death record includes the underlying cause of death (i.e. the medical condition judged to be the underlying cause according to the rules of the *10th Revision of the International Classification of Diseases*) and up to 15 medical conditions mentioned on the death certificate. Data is available from February 2019 onwards.

### Study population

#### Inclusion and exclusion criteria

We will include men and women alive on the threshold date, registered with an English GP using TPP software for at least 90 days, with known region, Index of Multiple Deprivation (IMD) and age information. For aim 1 (Zostavax), we will include people born between 1 September 1920 and 1 September 1948 (i.e., people between 65 and 93 years of age at the threshold date). For aim 2 (Shingrix), we will include people born between 1 September 1948 and 1 September 1968 (i.e., people between 55 and 75 years of age at the threshold date).

We will exclude people who received the vaccine under consideration (aim 1: Zostavax; aim 2: Shingrix) before the threshold date (1 September 2013/23). We will also exclude people with a diagnosis of shingles in the 28 days before the threshold date. (6–8)

We will exclude people with a diagnosis of dementia before the index date (1 February 2013 for Zostavax/to be determined for Shingrix).

For aim 1 (Zostavax), we will exclude people who have a known primary or acquired immunodeficiency state or patients who are receiving current immunosuppressive therapy, including high-dose corticosteroids, biological therapies or combination therapies, at the threshold date (1 September 2013) as they are not eligible to receive live vaccines such as Zostavax.

All OpenSAFELY studies exclude people who have registered as a Type 1 Opt-out. There are no recent data on the rate of Type 1 Opt-outs, but in December 2017, the Type 1 Opt-out rate was 3%. (9)

#### Follow-up

Follow-up will start on the index date and end at the earliest of the following dates: last data collection, outcome, deregistration, or death. Additionally, for aim 2 (Shingrix), follow-up will end at 5 years minus both the age bandwidth and the time between the threshold date and index date. For instance, if follow-up the chosen age bandwidth was +/- 2 years and follow-up started 0.5 years after the threshold date, we would stop follow-up at 2.5 years. This maximises follow-up while ensuring that the oldest people in our study population are censored when they become eligible for the Shingrix catch-up programme on their 70th birthday.

For this diagram, the study end date is 1 September 2025, and the age bandwidth is +/- 2 years. However, for our study, we will use data up to the last date of data collection with the MSE-optimal bandwidth approach, as specified below, to determine the age bandwidth.

The dark grey shading represents the campaign year under consideration (2013/2014). The light grey shading represents the age bandwidth.

Patients 5 and 8 are not eligible for the study as their 79th birthdays fall outside of the age bandwidth. Patient 6 has been excluded because they experienced the outcome before the start of follow-up. Among the remaining patients, we will compare those eligible for vaccination (green) with those not eligible for vaccination (blue).

For this diagram, the study end date is 1 September 2025, and the age bandwidth is +/- 2 years. However, for our study, we will use data up to the last date of data collection with the MSE-optimal bandwidth approach, as specified below, to determine the age bandwidth.

The dark grey shading represents the campaign year under consideration (2023/2024).

Patient 7 is not eligible for the study as their 65th birthday falls outside of the potential age bandwidth. Patients 2 and 4 have been excluded because they experienced the outcome before the start of follow-up.

Among the remaining patients, we will compare those eligible for vaccination (green) with those not eligible for vaccination (blue).

#### Study measures

Our study measures are specified below. The codelists to support these definitions are available (**Supplementary Table 2**).

#### Exposures

For aim 1, exposure is the one-dose live-attenuated HZ vaccine, Zostavax. We will define exposure date as the date of vaccination using vaccination records from primary care where VaccinationName is ‘Zostavax’ or ‘Shingles (Herpes Zoster) vaccine (live) powder and solvent for suspension for injection 0.65ml pfs’ (https://reports.opensafely.org/reports/opensafely-tpp-database-reference-values/#VaccinationReference-Table).

For aim 2, exposure is the two-dose recombinant subunit HZ vaccine, Shingrix. We will define exposure date as the date of the first vaccination using vaccination records from primary care where VaccinationName is ‘Shingrix’ or ‘Shingles (Herpes Zoster) adjuvanted rcmb vacc powder and suspension for suspension inj 0.5ml vials’ (https://reports.opensafely.org/reports/opensafely-tpp-database-reference-values/#VaccinationReference-Table).

#### Outcomes

The primary outcomes are dementia, shingles, postherpetic neuralgia and all-cause mortality. We will also study Alzheimer’s disease and vascular dementia, the most common dementia subtypes, as secondary outcomes. We will define our outcomes using the first date of diagnosis recorded in primary care after the index date, the first hospital admission after the index date with the diagnosis coded in any position, or a death record with the diagnosis listed as a primary or contributory cause of death. Note: data from hospital admissions and the death registry are only available from 2016 and 2019 onwards, respectively.

### Statistical analyses

#### Disclosure control

To prevent disclosure, all event counts will be rounded to the nearest 5 when reported, with risks calculated from rounded counts. Unrounded counts and risks will be used in analyses. Date of birth is captured as the month and year of birth.

#### Balance of dementia diagnoses at index

As stated above, we will exclude people with a diagnosis of dementia before the index date (1 February 2013 for Zostavax/to be determined for Shingrix). Before we implement this exclusion, we will quantify the number of dementia diagnoses in those eligible and not eligible for vaccination to examine whether they are balanced.

#### Determining the age range around the threshold

We will use a mean-squared error (MSE)-optimal procedure to determine the age range around the threshold, implemented using the R package ‘rdrobust’.

#### Preliminary analyses for aim 1 (Zostavax)

We will examine the cumulative coverage of Zostavax by age in months up to one year after the threshold date (1 September 2013) to confirm that the population captured in OpenSAFELY-TPP reflects the general population, which we used to pre-specify an index date of 1 February 2014. We will confirm a discontinuity is present by examining the vaccine coverage at the index date by age in monthly intervals.

#### Preliminary analyses for aim 2 (Shingrix)

We will conduct a series of preliminary analyses that do not include any outcome data to determine the minimum age for inclusion in the study on the threshold date (1 September 2023) and the index date when follow-up for the study will start. These analyses will include determining vaccine coverage after the threshold date (1 September 2023) and exploring the time taken following a person’s 65th birthday to be vaccinated. We will estimate the size of the discontinuity by examining vaccine coverage at the chosen index date by age in monthly intervals.

#### Incidence of dementia

We will examine the cumulative incidence of dementia by vaccine eligibility from the threshold date (1 September 2013/2023).

#### Vaccination coding

There are several other vaccines in the varicella-zoster vaccine family, including the chickenpox vaccine, which the NHS does not routinely offer to adults (TargetDisease=‘VARICELLA-ZOSTER’, https://reports.opensafely.org/reports/opensafely-tpp-database-reference-values/#VaccinationReference-Table). We will examine the cumulative coverage of these other vaccines by age in months up to one year after the threshold date (1 September 2013). In the event of non-trivial use of these vaccines, we will propose a protocol amendment detailing how we will account for use of these vaccines in our analyses.

#### Covariate balance

The key assumptions of regression discontinuity analyses are ‘continuity’ (that the risk of the outcome changes smoothly at the threshold in the absence of the intervention) and ‘exchangeability’ (that the distribution of confounders is similar just below and above the threshold). (10) To explore the plausibility of these assumptions, we will plot the distribution of the following characteristics by age at the threshold date in monthly intervals using data from primary care and admitted patient care in any position: sex (male, female); IMD quintiles; ethnicity (White, Mixed, Asian/Asian British, Black/Black British, Other, Unknown); practice region (East, East Midlands, London, North East, North West, South East, South West, Yorkshire and The Humber); previous clinical diagnoses (shingles; lower respiratory tract infection; asthma; atrial fibrillation; coronary heart disease; chronic kidney disease; chronic obstructive pulmonary disease; depression; diabetes mellitus; epilepsy; heart failure; hypothyroidism; serious mental illness; obesity; osteoporosis; peripheral arterial disease; rheumatoid arthritis; and stroke/TIA); lifestyle factors (smoking); and uptake of preventive health measures in the past 5 years (influenza vaccination, pneumococcal vaccination, statin use, antihypertensive use). We have decided not to plot the distribution of previous clinical diagnoses of cancer, learning disabilities and palliative care, and we will consider antihypertensive use in the past 5 years instead of a previous clinical diagnosis of hypertension, because these clinical diagnoses are not always well-captured in primary care data.

#### Sharp regression discontinuity

We will examine the effect of vaccine eligibility on each outcome *Y* by fitting the following regression model:

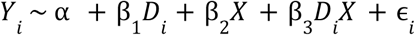

where, for person *i*, *Y* is a binary indicator for the outcome, *D* is a binary indicator for vaccine eligibility (ever/never), and *X* is the running variable (see Study Type).

We will express estimated effects as risk differences with associated 95% confidence intervals (CIs). The estimate of β_1_ represents the estimated difference in the outcome risk comparing people just above the threshold (who were targeted by the campaign) with people just below the threshold (who were not targeted by the campaign). A negative value means that the vaccination campaign lowers the rate of the outcome.

#### Fuzzy regression discontinuity

We will examine the effect of vaccination on each outcome using an instrumental variable approach, with vaccine eligibility as the instrument for vaccination. This will provide an estimate of the “local average treatment effect” (LATE) or “complier average causal effect” (CACE) at the threshold. This is the effect of vaccination among the subset of the population who are vaccinated only when eligible (the “compilers”).

For valid estimation, the three instrumental variable assumptions must hold:

- Relevance: The instrument must be associated with the intervention.
- Independence: The instrument and the outcome must have no uncontrolled common causes.
- Exclusion restriction: The instrument must only affect the outcome through the intervention. Additionally, we will make a fourth point-identifying assumption of ‘monotonicity’, that there are no “defiers” (people who are vaccinated only when ineligible). We will test the relevance assumption. The other assumptions are untestable but plausible.

We will implement two-stage least squares estimation. In the first stage, we will predict vaccine coverage by fitting the following regression model:

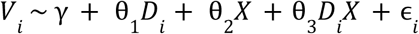

where, for person *i*, *V* is a binary indicator for vaccination, D is a binary indicator for vaccine eligibility, and *X* is the running variable (see Study Type).

In the second stage, we will examine the effect of vaccination on each outcome *Y* by fitting the following regression model:

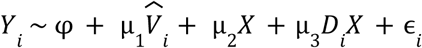

where, for person *i*, *Y* is a binary indicator for outcome, *V* is the predicted vaccine coverage from the first stage, and *X* is the running variable (see Study Type).

We will express estimated effects as risk differences with associated 95% CIs. The resulting estimate µ_1_ will represent the difference in the outcome risk among “compliers” (the subset of the population who are vaccinated only when eligible).

#### Estimation

We will use the R package ‘rdrobust’ to implement local linear regression (order of the local-polynomial, p = 1) with a triangular kernel weighting and heteroskedasticity-robust (HC0) standard errors for our estimation. (11) We will report both the conventional and robust bias-corrected estimates of the coefficient and 95% confidence intervals. The R code to implement these analyses is provided below for both the sharp and fuzzy regression discontinuity analyses.

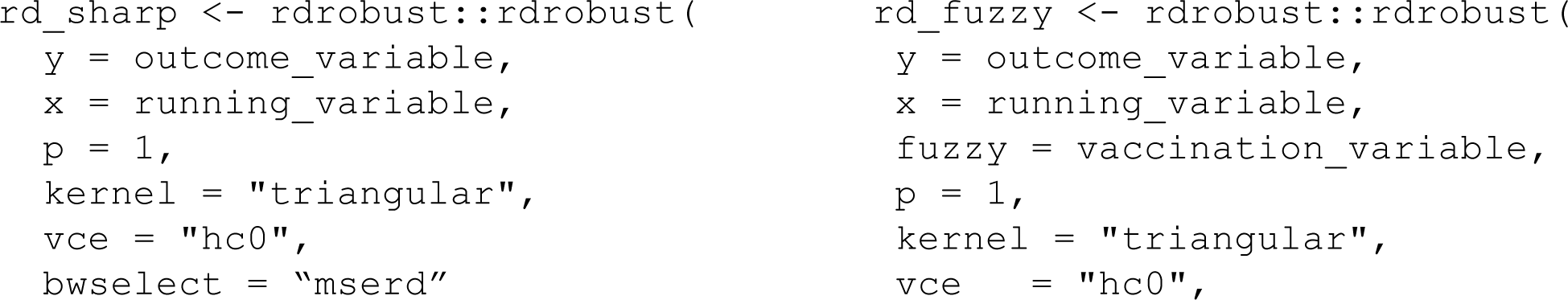
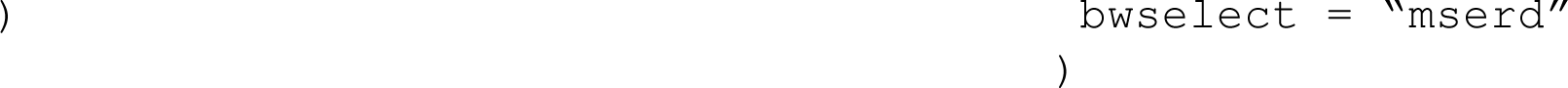

#### Sensitivity analyses

We will test the sensitivity of our primary analyses to different bandwidths:

- **Alternative MSE-optimal bandwidths.** We will repeat the analysis using a 0.5 and 2 times the MSE-optimal bandwidth to examine the degree to which our point estimates vary across different age bandwidths.
- **Fixed age bandwidths.** We will repeat the analysis using fixed age bandwidths of 1 and 2 years, in place of the MSE-optimal bandwidth.
- the MSE-optimal bandwidth for the outcome under consideration, we will test the sensitivity of our primary analyses to:
- **Imprecision of birth date.** We will repeat the analysis excluding people born in the month of the threshold date due to the imprecision in the recorded birth date.
- **Outcome codelist.** We will repeat the analysis for the outcome dementia, restricting our code list to codes that are also on the SNOMED CT mapped Charlson Comorbidity code list for dementia from Fortin et al. (12)
- **Exclusion criteria.** For aim 1 (Zostavax), we will repeat the sharp regression discontinuity analysis without the following exclusion criteria: ‘We will exclude people who received the vaccine under consideration before the threshold date (1 September 2013)’ and ‘We will exclude people with a diagnosis of dementia before the index date (1 February 2013).’

- **Threshold year.** Without these exclusions applied, we will also repeat the sharp regression discontinuity analysis at threshold years that are 3 years before and after the threshold year of interest. That is, instead of a threshold date of 1 September 2013, we will use threshold dates of 1 September 2010 and 1 September 2016. This will allow us to examine whether any observed effects are specific to the threshold year of interest, while maintaining the same age threshold. This sensitivity analysis must be conducted without the exclusion criteria applied due to the potential for differential exclusions in those who did and did not get vaccinated as part of the 2013 programme in analyses that use a later threshold date.
- **Choice of outcome.** We will examine whether observed effects are specific to dementia by repeating the analysis for selected outcomes associated with morbidity and mortality that were required to have patient registers as part of the Quality and Outcomes Framework (QOF) in 2013/14. The outcomes we will look at are: asthma; atrial fibrillation; coronary heart disease; chronic kidney disease; chronic obstructive pulmonary disease; depression; diabetes mellitus; epilepsy; heart failure; hypothyroidism; serious mental illness; obesity; osteoporosis; peripheral arterial disease; rheumatoid arthritis; stroke and TIA. Negative control outcomes are those the instrument should not plausibly affect; thus, an observed association between the instrument and a negative control outcome suggests a violation of key IV assumptions, such as independence or the exclusion restriction. While they cannot prove the assumptions hold when no association is found, they provide a useful falsification test that can reveal otherwise hidden biases. It is difficult to identify true negative control outcomes; hence, we have included a range of potential options.
- **Annual incremental follow-up.** We will repeat our analyses limiting follow-up to annual increments (e.g., up to 1 year, up to 2 years, …) until the maximum length of follow-up, which serves as our primary analysis. This will include analyses limited to 7 and 8 years of follow-up in line with the previous Welsh analysis.
- **Restriction to primary care.** Data from hospital admissions and the death registry are only available from 2016 and 2019 onwards, respectively. For aim 1 (Zostavax), we will repeat our analyses using outcomes defined in primary care only, as this reflects data with complete coverage during our study period.
- **Kernel function used to construct the local polynomial estimator.** We will repeat our analyses using a uniform kernel function in place of the triangular kernel function specified for our main analysis.
- **Order of the local polynomial estimator.** We will repeat our analyses using local quadratic regression in place of the local linear regression specified for our main analysis.

#### Subgroup analyses

We will perform the following subgroup analyses for all vaccines:

- Sex (male/female)
- Evidence of cognitive impairment at threshold date

If feasible, for aim 2 (Shingrix), we will also perform the following subgroup analyses:

- Previous receipt of Zostavax (yes/no)

#### Limitations

Our study is subject to the following limitations:

- **Generalizability.** Findings from this study apply to people who are within the age bandwidth around the threshold age, so may not extend to substantially younger or older populations.
- **Exchangeability.** Regression discontinuity relies on the assumption that confounding variables among people just above and below the threshold are expected to be similar. We will explore the plausibility of this assumption by plotting the distribution of a range of relevant characteristics by age in 3-month intervals.
- **Date of outcome ascertainment.** Dementia develops gradually and is often present for some time before it is recognised and recorded in routine healthcare data. The recorded diagnosis date therefore reflects the time of clinical ascertainment rather than disease onset.
- **Diagnostic coding accuracy and consistency.** Diagnostic practices, coding accuracy, and the timeliness of recording may have changed over the study period, potentially leading to misclassification and temporal inconsistencies in outcome ascertainment.

#### Disclosure control

To manage the risk of disclosure–that is, the reidentification of people within the data– only aggregated results will be viewed and requested for release from the secure environment, and statistical disclosure control (SDC) will be applied to all results, as detailed in <u>Statistical methods / Disclosure control</u>. This will include redaction of low values, rounding, and/or redesigning outputs to avoid sparse table cells, as required. To ensure SDC has been properly applied, all results will be reviewed by two trained output checkers before release from the secure environment.

#### Software and reproducibility

Data management will be performed using Python, with analyses carried out using R. Code for data management and analysis, as well as code lists, will be archived online using GitHub: https://github.com/opensafely/zostavax-dementia.

### Patient and Public Involvement and Engagement (PPIE)

#### Project-specific PPIE

Following the study on the effect of the shingles vaccine in Wales [2], members of that team (PG) solicited the input of a Personal and Public Involvement panel at the Aberdeen Centre for Health Data Science. This panel consisted of eight diverse lay members who discussed the findings of the prior work and the potential value of a subsequent study. After careful discussion and evaluation, the panel considered the continuation of this type of research meaningful and beneficial.

#### OpenSAFELY PPIE

OpenSAFELY has involved patients and the public in various ways: we developed a public website that provides a detailed description of the platform in language suitable for a lay audience (https://opensafely.org); we have participated in two citizen juries exploring public trust in OpenSAFELY; we have co-developed an explainer video (https://www.opensafely.org/about/); we have patient representation who are experts by experience on our OpenSAFELY Oversight Board; we have partnered with Understanding Patient Data to produce lay explainers on the importance of large datasets for research; we have presented at various online public engagement events to key communities (e.g., Healthcare Excellence Through Technology; Faculty of Clinical Informatics annual conference; NHS Assembly; HDRUK symposium); and more. To ensure the patient voice is represented, we are working closely to decide on language choices with appropriate medical research charities (e.g., Association of Medical Research Charities). We will share information and interpretation of our findings through press releases, social media channels, and plain language summaries.

#### Dissemination

We plan to disseminate our findings via peer-reviewed scientific journal publications as well as presentations at relevant academic conferences (e.g., the Alzheimer’s Association International Conference). We will report our studies using the RECORD reporting guidelines. (13)

#### Timelines and approvals

Our OpenSAFELY project application was approved on 1 June 2026 (Project ID: POS-2026-3003): https://www.opensafely.org/project/pos-2026-3003/. Code will be developed against dummy data. We will commence running the project using real data following the posting of this protocol in the public domain. NHS England will oversee the final approval for all publication-ready papers, reports or presentations, to check that the outputs align with the stated application purpose.

#### Ethics

This research study has been reviewed by the Medical Sciences Interdivisional Research Ethics Committee (MS IDREC) at the University of Oxford in accordance with the University’s regulations and policy for ethics approval of research involving human participants, human tissue and/or personal data. The research study received a favourable opinion (Decision Date: 18 Mar 2026; MS IDREC ID: 3026906).

## Information governance

Information governance for projects conducted in OpenSAFELY can be found here: https://docs.opensafely.org/contributorship-and-content-guidance/#information-governance

## Data Availability

No data produced in the present work (protocol).

https://www.opensafely.org/

## Supplementary tables

**Supplementary table 1:**
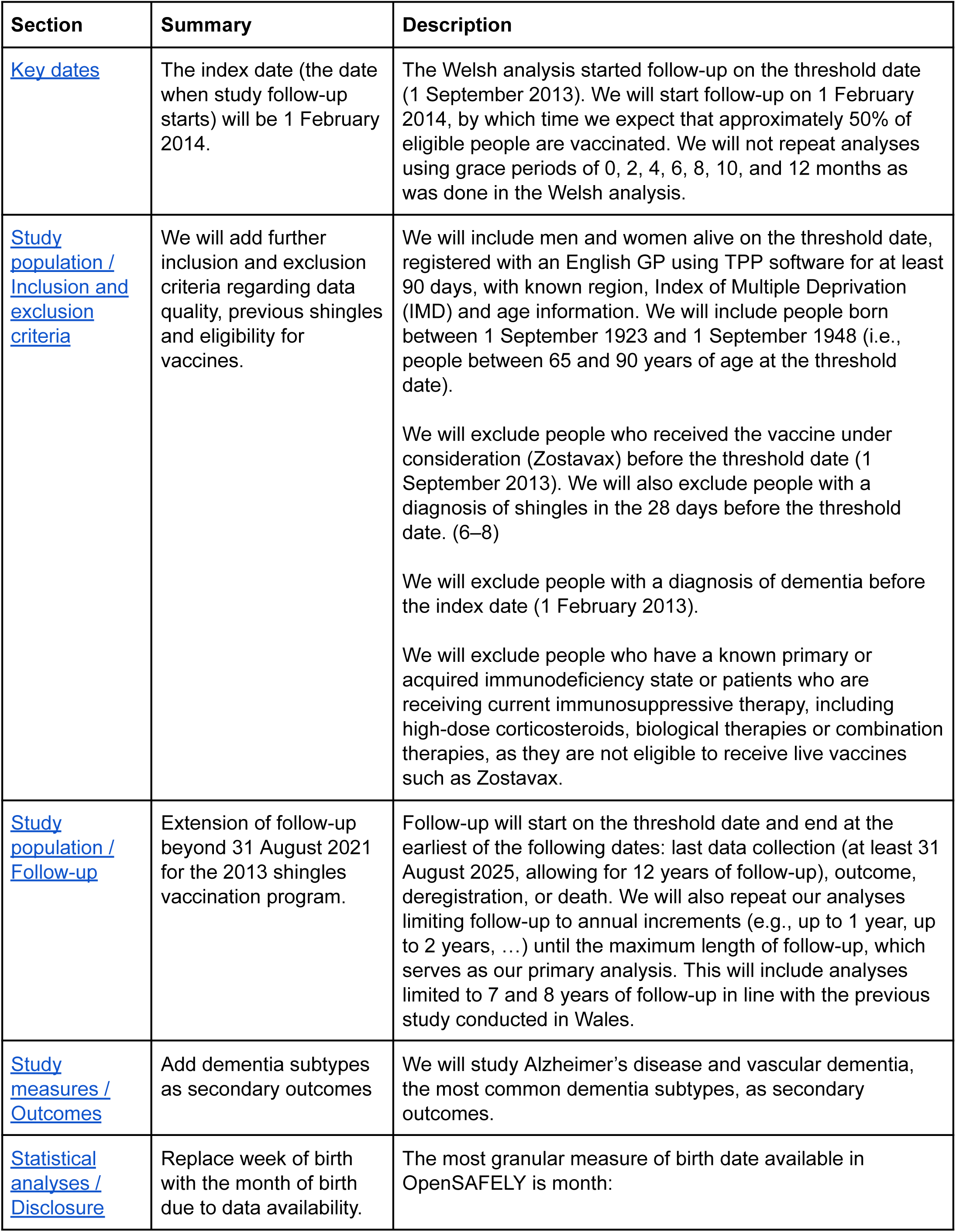
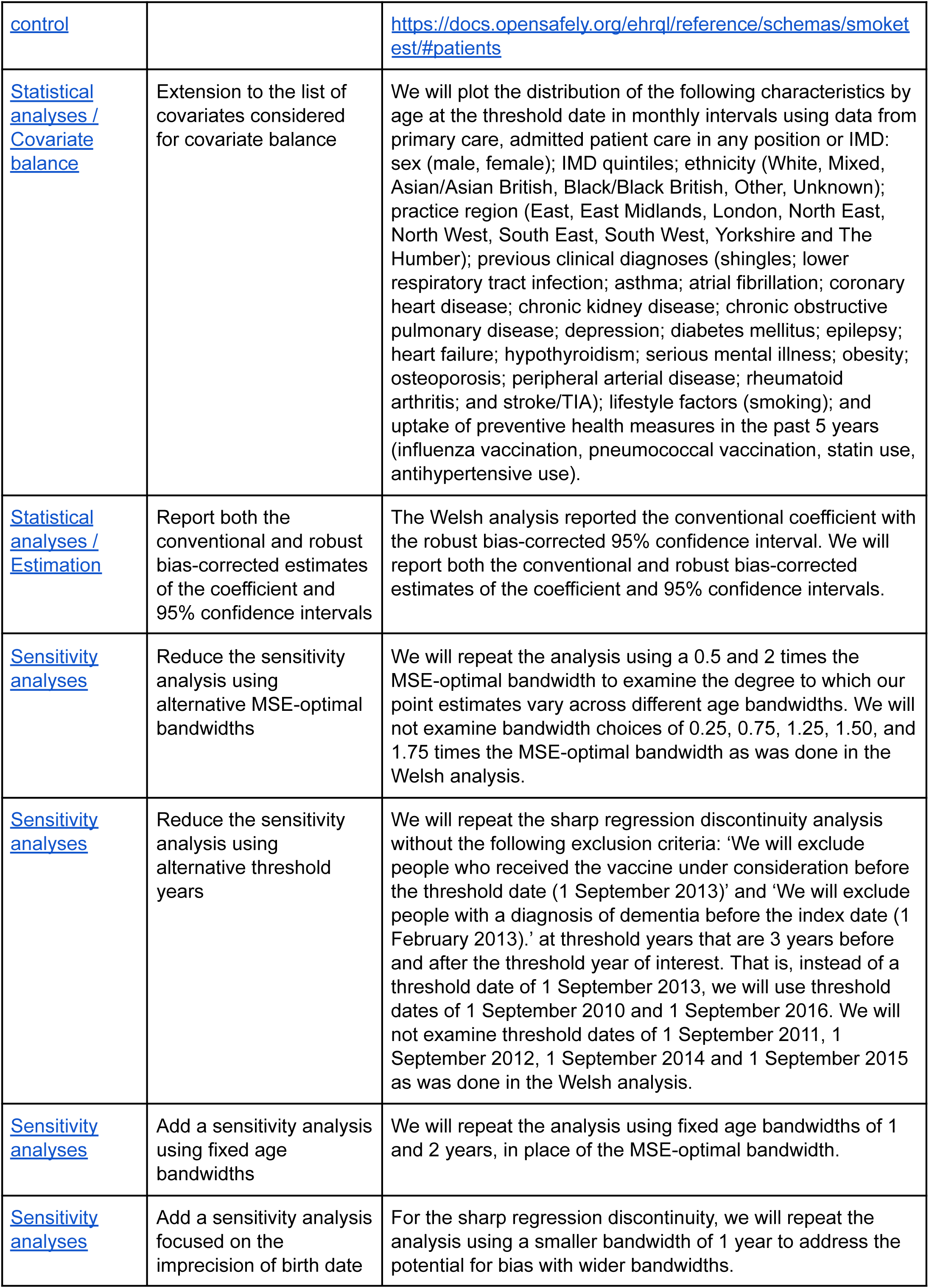
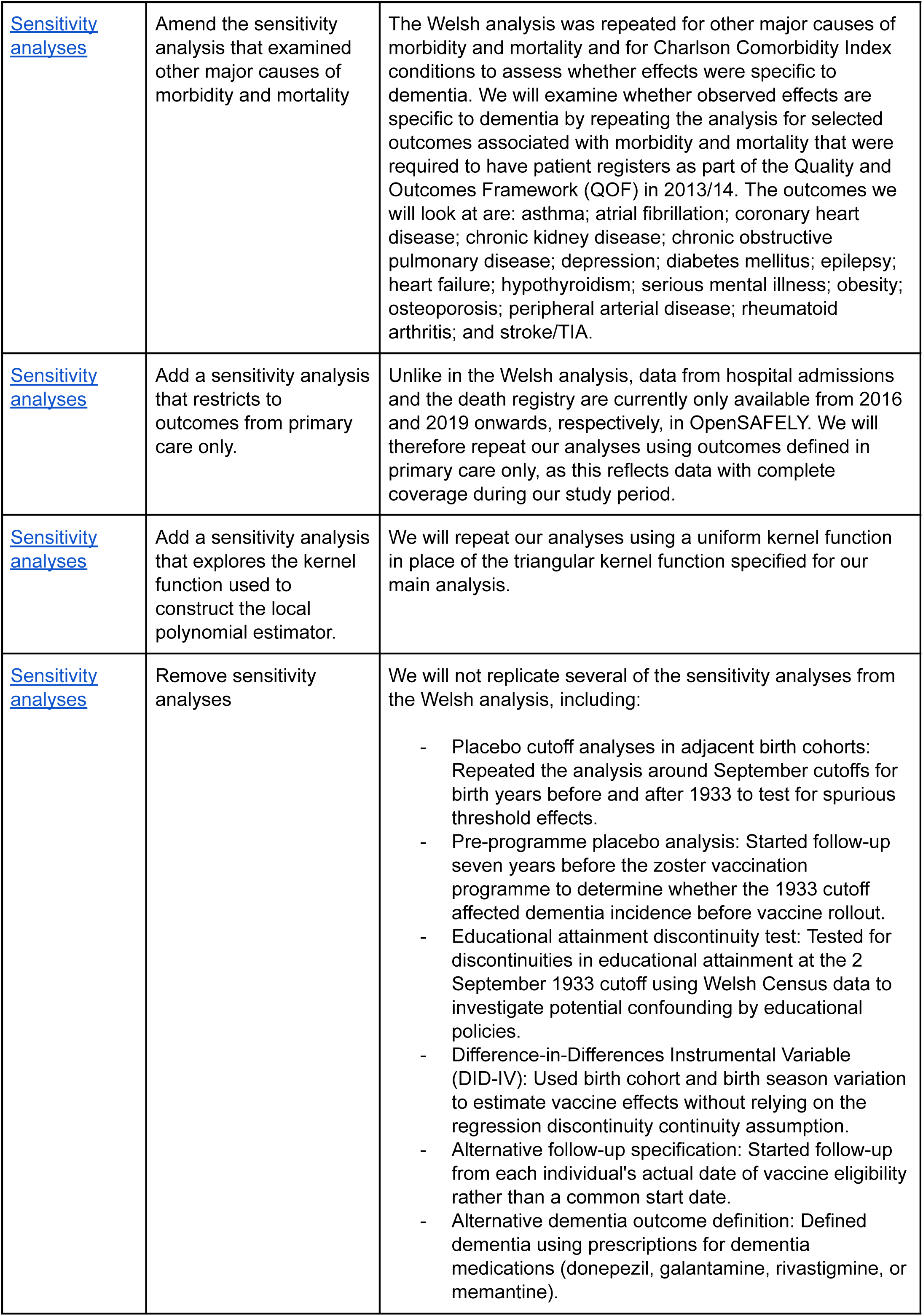
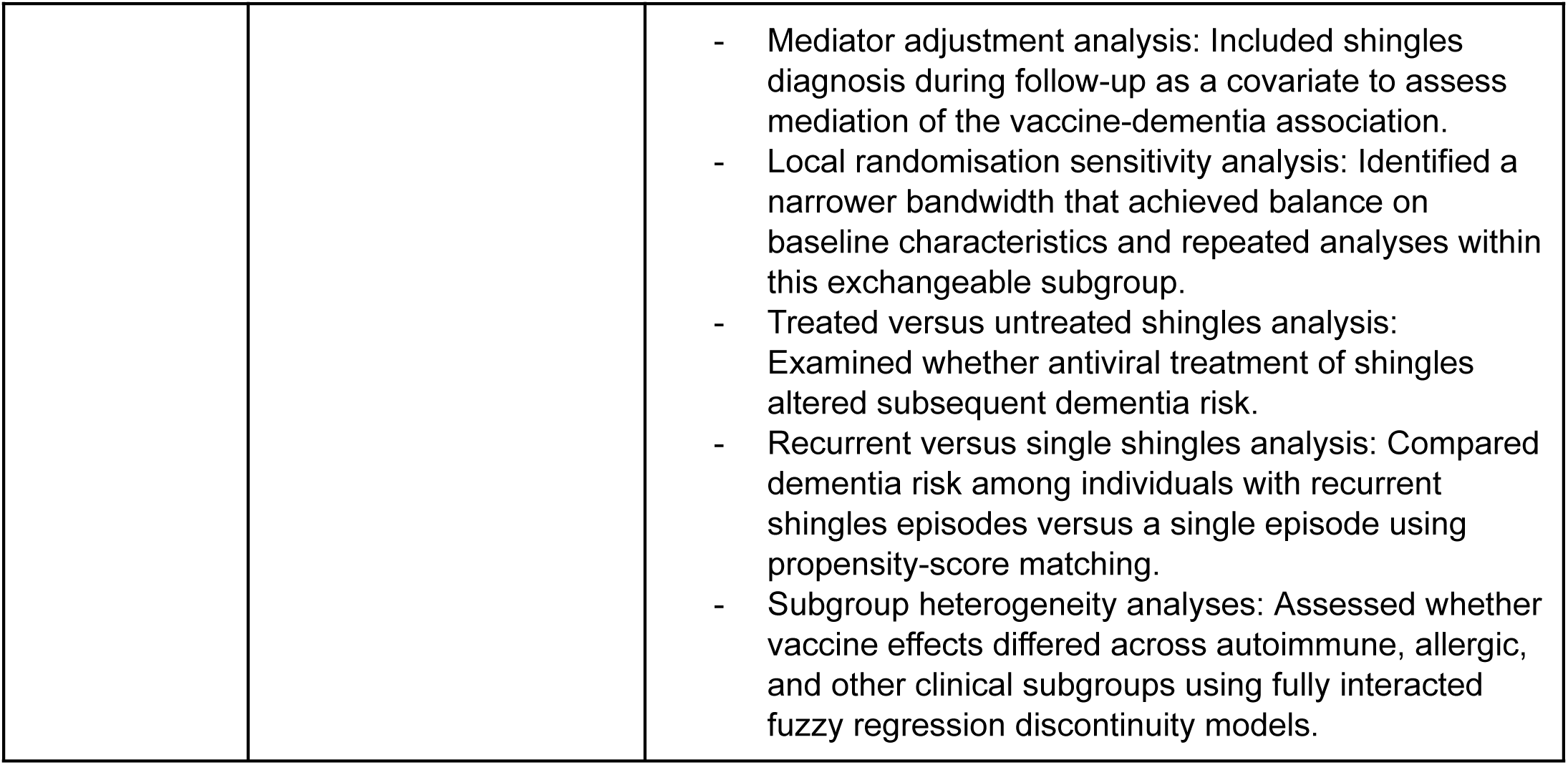
Deviations from Welsh analysis for aim 1 (Zostavax)

**Supplementary table 2:** Codelists.

